# Impact of the COVID-19 Vaccination Program on Case Incidence, Emergency Department Visits, and Hospital Admissions among Children Aged 5–17 Years during the Delta and Omicron Periods —United States, December 2020 to April 2022

**DOI:** 10.1101/2022.10.07.22280822

**Authors:** Katherine G Topf, Michael Sheppard, Grace E. Marx, Ryan E Wiegand, Ruth Link-Gelles, Alison M Binder, Andrea J Cool, B. Casey Lyons, Sohyun Park, Hannah E Fast, Arthur Presnetsov, G Roseric Azondekon, Karl A Soetebier, Jennifer Adjemian, Kamil E Barbour

**Author notes:** **Corresponding Author:** Kamil Elie Barbour, PhD, MPH, MS, 4770 Buford Highway, Atlanta, GA, 30341. **CDC Disclaimer:** The findings and conclusions in this report are those of the author(s) and do not necessarily represent the official position of the Centers for Disease Control and Prevention (CDC).

## Abstract

**Background:** In the United States, national ecological studies suggest a positive impact of COVID-19 vaccination coverage on outcomes in adults. However, the national impact of the vaccination program on COVID-19 in children remains unknown. To determine the association of COVID-19 vaccination with U.S. case incidence, emergency department visits, and hospital admissions for pediatric populations during the Delta and Omicron periods.

**Methods:** We conducted an ecological analysis among children aged 5–17 and compared incidence rate ratios (RRs) of COVID-19 cases, emergency department visits, and hospital admissions by pediatric vaccine coverage, with jurisdictions in the highest vaccine coverage quartile as the reference.

**Results:** RRs comparing states with lowest pediatric vaccination coverage to the highest pediatric vaccination coverage were 2.00 and 0.64 for cases, 2.96 and 1.11 for emergency department visits, and 2.76 and 1.01 for hospital admissions among all children during the Delta and Omicron periods, respectively. During the 3-week peak period of the Omicron wave, only children aged 12–15 and 16–17 years in the states with the lowest versus highest coverage, had a significantly higher rate of emergency department visits (RR=1.39 and RR=1.34, respectively).

**Conclusions:** COVID-19 vaccines were associated with lower case incidence, emergency department visits and hospital admissions among children during the Delta period but the association was weaker during the Omicron period. Pediatric COVID-19 vaccination should be promoted as part of a program to decrease COVID-19 impact among children; however, vaccine effectiveness may be limited when available vaccines do not match circulating viral variants.

## Introduction

In the United States, Pfizer-BioNTech was recommended on December 12, 2020, for children aged 16–17 years, May 12, 2021, for children aged 12–15 years, and November 2, 2021, for children aged 5–11 years^[1-3]^. As of May 6, 2022, 62.6% of children aged 16–17 years, 57.6% of children aged 12–15 years, and 28.8% of children aged 5–11 years were fully vaccinated with the Pfizer-BioNTech COVID-19 vaccination series^[4]^.

Although children are less likely to develop severe COVID-19 compared to adults, the pandemic has nonetheless had a serious direct and indirect negative impact on children^[5]^. Among children aged <18 years, 12,752,227 cases and 1,536 deaths due to COVID-19 have been reported as of May 16, 2022^[6]^. Children can also suffer delayed sequelae from COVID-19 including multisystem inflammatory syndrome which usually requires hospitalization and can result in disability and death^[7]^. In addition, isolation during infection and quarantine after exposure to sick contacts results in fewer days in school where children can access nutritious food and social and mental health support^[8]^.

Prior U.S. national studies of adult populations have shown that people living in areas where COVID-19 vaccination of adults is higher have lower case incidence, lower hospitalization rates, and fewer emergency department visits due to COVID-19^[9-10]^. With the introduction of pediatric COVID-19 vaccinations, it is important to assess this impact among children. Previous studies indicate vaccines are effective for the prevention of infection and hospitalization, though the effectiveness is lower during Omicron circulation than for previous variants^[11-13]^. We conducted an ecological study to determine the association of state-level COVID-19 vaccination rates with case incidence, emergency department visits, and hospital admissions in the pediatric population during two crucial periods of the COVID-19 pandemic when the Delta and Omicron variants predominated.

## Methods

### Study Design

To assess the impact of pediatric vaccination coverage on COVID-19 in children, we analyzed weekly time series data for COVID-19 cases, emergency department visits with diagnosed COVID-19, and hospital admissions of patients with lab-confirmed positive SARS-CoV-2 results from December 13, 2020, to April 30, 2022. We stratified data for COVID-19 cases and emergency department diagnoses by pediatric age groups for children aged 5–11, 12–15, and 16–17 years; we included aggregated hospital weekly admission data among children aged 0–17 years. We defined the analytic period to capture periods before and after COVID-19 vaccine was authorized for each pediatric age group. To compare outcomes in jurisdictions (U.S. states and the District of Columbia) in the lowest quartile for pediatric vaccination coverage to jurisdictions in the highest quartile for pediatric vaccination coverage, we calculated rate ratios with high coverage jurisdictions used as the reference group for: 1) all weeks included in the analytical period; and 2) date ranges aligning with predominance of the Delta and Omicron variants in the United States, and the 3-week peak periods of Delta and Omicron waves. We aggregated county-level vaccination coverage data into jurisdiction-level data and then grouped jurisdictions into vaccination coverage categories using quartiles for vaccination coverage rate estimates for children aged 5–17 years as of April 2022.

This activity was reviewed by the Centers for Disease Control and Prevention’s (CDC) Human Research Protection Office and determined to be exempt from human participants’ research regulations, including the need for documented written consent, as the activities involved identification, control or prevention of disease in response to an immediate public health threat. It was conducted consistent with applicable federal law and CDC policy (See e.g., 45 C.F.R. part 46, 21 C.F.R. part 56; 42 U.S.C. §241(d); 5 U.S.C. §552a; 44 U.S.C. §3501 et seq.).

### Data Sources

#### Cases

Jurisdictional (state (50), territorial (3) and local (2)) health departments voluntarily submit de-identified individual-level data for COVID-19 cases to CDC via the COVID-19 Case Report Form and the National Notifiable Diseases Surveillance System^[14-15]^. These data sources are aggregated for use as a single dataset. Age was available for 99% of recorded cases. In this analysis, we used data from 33 states, representing 70% of US population under age 18; we excluded jurisdictions if less than 80% of records contained residential county information, if less than 90% of records contained relevant date information, or if the jurisdiction was missing data for more than 1 week in the study period. Jurisdictions included were Alabama, Arizona, Arkansas, California, Colorado, Delaware, Georgia, Idaho, Illinois, Indiana, Kansas, Louisiana, Maine, Massachusetts, Minnesota, Missouri, Montana, Nevada, New Jersey, New Mexico, New York, North Carolina, North Dakota, Ohio, Oregon, Pennsylvania, South Carolina, Tennessee, Utah, Vermont, Virginia, Washington, and Wyoming.

#### Emergency Department Visits

We collected emergency department visit data from the National Syndromic Surveillance Program (NSSP), representing encounters with a diagnosis of COVID-19, defined by having any of the following discharge diagnoses: International Classification of Diseases, Tenth Revision codes U07.1 or J12.82 or Systematized Nomenclature of Medicine (SNOMED) codes 840539006, 84054404, or 840533007. NSSP collects electronic health data from 73% of non-federal hospital facilities across all jurisdictions^[16]^. We applied data quality filters to only include data from facilities with high completeness of discharge diagnoses (average weekly completeness ≥75%) and consistent reporting over the analytic period. Fewer than 50% of facilities in California, Hawaii, Iowa, Minnesota, and Oklahoma send data to the NSSP; we excluded emergency department visit data from these jurisdictions from this analysis. Additionally, we excluded jurisdictions with low completeness for discharge diagnoses (Missouri) and those with missing data during the analytical period (Maryland).

#### Hospital Admissions

De-identified COVID-19 hospitalization data are reported by over 6,000 hospitals in the United States through the U.S. Department of Health and Human Services Unified Hospital Data Surveillance System (UHDSS), which collects daily hospital-level data from all hospitals registered with the Centers for Medicare and Medicaid Services (CMS) as of June 1, 2020, and data from hospitals not registered with CMS but reporting since July 15, 2020^[17]^. Facility subtype designations are determined by CMS. We excluded psychiatric, rehabilitation, and religious non-medical facilities from analyses, because these facilities are required to report per conditions of participation, but do not typically treat or admit patients with acute COVID-19. Data included in analyses represent all jurisdictions and were both consistent and complete during the analytic period (at least 5146 [98%] of 5251 hospitals meeting inclusion criteria reported COVID-19 admissions data on any given day between December 13, 2020, and April 30, 2022). New admissions of pediatric patients aged 0 – 17 years with confirmed COVID-19 obtained from UHDSS were available as aggregate counts reported daily during the study period. Reporting of new admissions of patients with confirmed COVID-19 by narrower age ranges (0–4, 5–11, and 12–17 years) began in February 2022. Thus, we conducted analyses of pediatric hospital admissions data for children aged 0–17 years; further stratification by age was not possible due to limited availability of the more granular age-specific admissions data.

#### Vaccine Administration Data

Individual-level COVID-19 vaccine administration data are submitted to CDC via jurisdictional immunization information systems, the Vaccine Administration Management System, or direct data submission^[18-19]^. The data are de-duplicated, de-identified, and subjected to systematic quality control checks prior to analysis^[20]^. For this analysis, we included Pfizer-BioNTech vaccines administered to children aged 5–17 years by April 30, 2022, in all reporting jurisdictions.

### Statistical Analysis

We used U.S. 2019 population estimates to calculate jurisdiction pediatric vaccination coverage rates and cases and hospital admission rates per 100,000 population^[21]^. We calculated jurisdiction pediatric vaccination coverage quartiles using cumulative percentages for children aged 5–17 years who had completed the COVID-19 vaccination series. To estimate rate ratios comparing jurisdictions with lower vaccination coverage (lowest, second lowest, and second highest vaccination coverage quartiles) to jurisdictions in the highest vaccination coverage quartile, we used Poisson regression models and generalized estimating equations (GEE) to account for clustering of data by state. We calculated 95% confidence intervals (CIs) using robust standard errors.

We calculated ratios for emergency department visits using the total number of visits rather than population-based estimates. To assess differences in lower and highest vaccination coverage jurisdictions over time, we calculated rate ratios for case counts, emergency department visits, and hospital admissions for: 1) all weeks from December 13, 2020, to April 30, 2022; 2) time periods aligning with predominance of the Delta and Omicron variants in the United States (Delta: June 20, 2021–October 31, 2021; Omicron: December 19, 2021–April 30, 2022); and 3) 3-week time periods aligning with peaks in case incidence during the Delta and Omicron variant time periods (Delta: August 22, 2021–September 11, 2021; Omicron: January 2, 2022–January 22, 2022). We calculated jurisdiction coverage quartiles separately for the Delta and Omicron time periods, using coverage estimates from the mid-point of each period (Delta: September 5, 2021; Omicron: February 20, 2022). For weekly rate ratio calculations, we used coverage estimates as of April 30, 2022 to determine jurisdictions in the lowest and highest (reference) coverage quartiles. We kept jurisdictions in the lowest and highest coverage quartiles fixed over the analytic period for weekly rate ratio calculations. For COVID-19 cases and emergency department visits, we calculated rate ratios separately for children aged 5–11, 12–15, and 16–17 years, and for children aged 5–17 years overall. Rate ratio calculations for hospital admissions were limited to children aged 0–17 years. We used R version 4.3.1 for all statistical analyses.

## Results

As of April 30, 2022, vaccination coverage remained highest for children aged 16–17 years (62.8%), followed by children aged 12–15 (56.7%) and 5–11 (27.3%) years (Figure 1). For the age groups of 5–11 and 12–15 years, the increase in vaccination coverage was greatest in the first 1–2 months following FDA Emergency Use Authorization and CDC recommendation of the COVID-19 and slowed dramatically in subsequent months.

**Figure 1:**
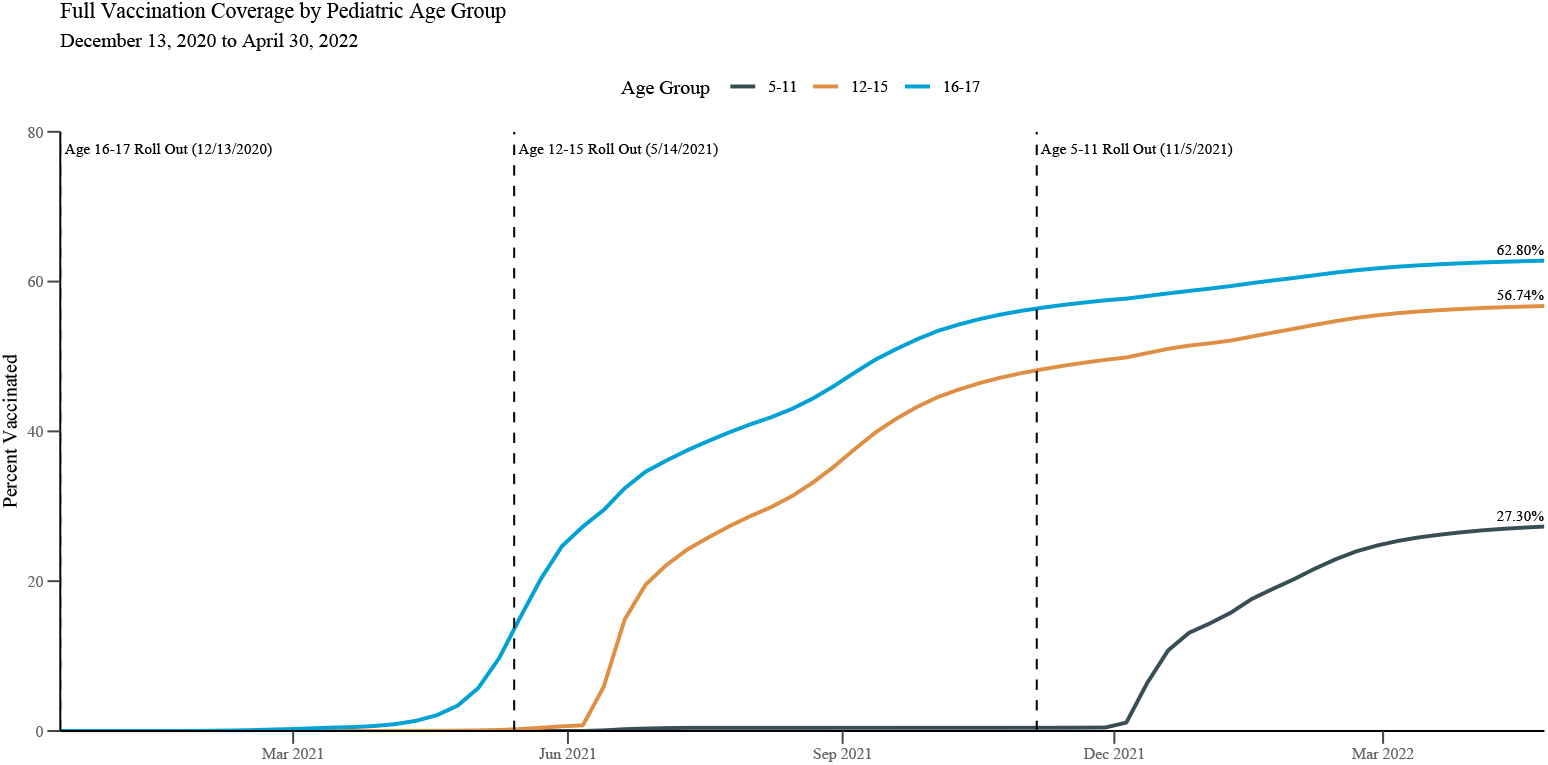
Weekly “fully vaccinated” pediatric vaccination coverage rates by age group among children aged 5–17 years, U.S., from December 13, 2020, through April 30, 2022

During the Delta and Omicron periods of predominance in the US, case incidence and the percentage of emergency department visits with diagnosed COVID-19 in children aged 5–17 years were 4,206 per 100,000 persons (cases) and 3.24% (emergency department visits) during the Delta period, and 8,724 per 100,000 persons (cases) and 3.81% (emergency department visits) during the Omicron period. Hospital admission rates in children aged 0–17 years during the Delta and Omicron periods were 37 per 100,000 persons and 67 per 100,000 persons, respectively (Appendix table 1).

In the overall pediatric population, rate ratios comparing jurisdictions in the lowest pediatric vaccination coverage quartile (8%–12% for cases and ED visits, 8%–13% for hospital admissions) (Table 2) to the highest pediatric vaccination coverage quartile (22%–28% for cases, 22%–29% for ED visits and hospital admissions) during the period of Delta predominance were 2.00 (95% CI: 1.50–2.67) for cases, 2.96 (95% CI: 2.13–4.11) for emergency department visits, and 2.76 (95% CI: 2.04–3.74) for hospital admissions (Table 1, Figure 2). In contrast, during the Omicron period of predominance, low coverage jurisdictions (19%–27%) relative to high coverage jurisdictions (46%–56%) were associated with lower case incidence: (0.64 (95% CI: 0.45–0.91)). Vaccination coverage was not associated with emergency department visits (1.11 (95% CI: 0.85– 1.44)) and hospital admissions (1.01 (95% CI: 0.80–1.28)) during the Omicron period of predominance (Table 1).

**Table 1.** Rate ratios among children aged 5–17 years by outcome comparing COVID-19 pediatric vaccination coverage quartiles of states to states in the highest vaccination coverage quartile during Delta and Omicron predominant periods – by age group and overall

| Outcome | State | Rate Ratio (95% CIs) |  |  |  |  |  |  |  |  |  |  |  |  |  |  |  |
| --- | --- | --- | --- | --- | --- | --- | --- | --- | --- | --- | --- | --- | --- | --- | --- | --- | --- |
|  | Vaccination | 5–11 |  |  |  | 12–15 |  |  |  | 16–17 |  |  |  | All<sup>a</sup> |  |  |  |
|  | Coverage | Delta |  | Omicron |  | Delta |  | Omicron |  | Delta |  | Omicron |  | Delta |  | Omicron |  |
|  | Quartile | Full<sup>b</sup> | Peak<sup>c</sup> | Full<sup>d</sup> | Peak<sup>e</sup> | Full | Peak | Full | Peak | Full | Peak | Full | Peak | Full | Peak | Full | Peak |
| Cases<sup>f</sup> | Highest | Reference |  |  |  |  |  |  |  |  |  |  |  |  |  |  |  |
|  | Second Highest | 1.25<br>(0.99,<br>1.58) | 1.14<br>(0.82,<br>1.58) | 0.65<br>(0.49,<br>0.85) | 0.63<br>(0.43,<br>0.92) | 1.24<br>(0.98,<br>1.56) | 1.16<br>(0.81,<br>1.66) | 0.72<br>(0.55,<br>0.94) | 0.73<br>(0.49,<br>1.09) | 1.26<br>(1.01,<br>1.58) | 1.24<br>(0.91,<br>1.69) | 0.78<br>(0.59,<br>1.03) | 0.79<br>(0.52,<br>1.23) | 1.25<br>(0.99,<br>1.57) | 1.16<br>(0.83,<br>1.62) | 0.69<br>(0.53,<br>0.91) | 0.69<br>(0.46,<br>1.02) |
|  | Second Lowest | 1.54<br>(1.24,<br>1.91) | 2.05<br>(1.44,<br>2.91) | 0.71<br>(0.48,<br>1.03) | 0.77<br>(0.49,<br>1.23) | 1.85<br>(1.49,<br>2.30) | 2.60<br>(1.82,<br>3.72) | 0.80<br>(0.57,<br>1.13) | 0.92<br>(0.59,<br>1.45) | 1.84<br>(1.49,<br>2.28) | 2.48<br>(1.80,<br>3.43) | 0.84<br>(0.61,<br>1.16) | 0.96<br>(0.61,<br>1.51) | 1.68<br>(1.36,<br>2.08) | 2.29<br>(1.62,<br>3.23) | 0.76<br>(0.54,<br>1.08) | 0.85<br>(0.54,<br>1.34) |
|  | Lowest | 1.78<br>(1.34,<br>2.37) | 2.78<br>(1.86,<br>4.15) | 0.61<br>(0.42,<br>0.88) | 0.60<br>(0.37,<br>0.96) | 2.30<br>(1.71,<br>3.08) | 3.78<br>(2.46,<br>5.83) | 0.66<br>(0.47,<br>0.94) | 0.67<br>(0.42,<br>1.07) | 2.18<br>(1.61,<br>2.95) | 3.40<br>(2.22,<br>5.21) | 0.69<br>(0.48,<br>0.97) | 0.70<br>(0.43,<br>1.12) | 2.00<br>(1.50,<br>2.67) | 3.19<br>(2.11,<br>4.82) | 0.64<br>(0.45,<br>0.91) | 0.64<br>(0.40,<br>1.02) |
| Emergency | Highest | Reference |  |  |  |  |  |  |  |  |  |  |  |  |  |  |  |
| Department Visits<sup>g</sup> | Second Highest | 1.83<br>(1.13, 2.97) | 2.09<br>(1.19, 3.65) | 0.94<br>(0.75, 1.17) | 1.07<br>(0.81, 1.42) | 2.06<br>(1.22, 3.50) | 2.60<br>(1.43, 4.72) | 1.05<br>(0.85, 1.29) | 1.20<br>(0.91, 1.60) | 1.87<br>(1.10, 3.19) | 2.02<br>(1.06, 3.84) | 1.04<br>(0.84, 1.29) | 1.18<br>(0.88, 1.58) | 1.90<br>(1.15, 3.15) | 2.21<br>(1.23, 3.97) | 0.99<br>(0.80, 1.22) | 1.14<br>(0.86, 1.50) |
|  | Second Lowest | 2.16<br>(1.41, 3.30) | 2.25<br>(1.45, 3.52) | 1.02<br>(0.78, 1.33) | 1.14<br>(0.86, 1.53) | 2.59<br>(1.77, 3.80) | 2.84<br>(1.88, 4.29) | 1.16<br>(0.91, 1.48) | 1.32<br>(1.00, 1.74) | 2.41<br>(1.68, 3.45) | 2.41<br>(1.60, 3.62) | 1.13<br>(0.89, 1.43) | 1.26<br>(0.95, 1.68) | 2.34<br>(1.59, 3.44) | 2.45<br>(1.62, 3.70) | 1.08<br>(0.84, 1.39) | 1.22<br>(0.92, 1.62) |
|  | Lowest | 2.68<br>(1.89, 3.82) | 2.95<br>(2.09, 4.18) | 1.03<br>(0.77, 1.37) | 1.19<br>(0.87, 1.64) | 3.30<br>(2.38, 4.58) | 4.07<br>(2.89, 5.74) | 1.19<br>(0.92, 1.54) | 1.39<br>(1.02, 1.90) | 3.08<br>(2.28, 4.17) | 3.46<br>(2.38, 5.02) | 1.18<br>(0.93, 1.50) | 1.34<br>(1.00, 1.78) | 2.96<br>(2.13, 4.11) | 3.41<br>(2.41, 4.82) | 1.11<br>(0.85, 1.44) | 1.28<br>(0.95, 1.74) |
| Hospital Admission<sup>h</sup> | Highest | Reference |  |  |  |  |  |  |  |  |  |  |  |  |  |  |  |
|  | Second Highest |  |  |  |  |  |  |  |  |  |  |  |  | 1.97<br>(1.27, 3.06) | 2.25<br>(1.23, 4.10) | 1.30<br>(1.00, 1.69) | 1.22<br>(0.91, 1.64) |
|  | Second Lowest |  |  |  |  |  |  |  |  |  |  |  |  | 2.67<br>(1.61, 4.44) | 3.04<br>(1.60, 5.77) | 1.36<br>(0.98, 1.89) | 1.19<br>(0.87, 1.63) |
|  | Lowest |  |  |  |  |  |  |  |  |  |  |  |  | 2.76<br>(2.04, 3.74) | 3.73<br>(2.70, 5.14) | 1.01<br>(0.80, 1.28) | 1.04<br>(0.81, 1.34) |

<sup>a</sup> Hospital admission data are only available in aggregate for children aged 0–17 years during these periods. For cases and ED visits, data are shown for pediatric patients aged 5–17 years. <sup>b</sup> Full period of Delta predominance defined as June 20, 2021 through October 31, 2021 <sup>c</sup> Peak period of Delta predominance defined as August 22, 2021 through September 11, 2021 <sup>d</sup> Full period of Omicron predominance defined as December 19, 2021 through April 30, 2022 <sup>e</sup> Peak period of Omicron predominance defined as January 2, 2022 through January 22, 2022 <sup>f</sup> Jurisdictions excluded from case data source include Alaska, Connecticut, District of Columbia, Florida, Kentucky, Hawaii, Iowa, Maryland, Missouri, Mississippi, Nebraska, New Hampshire, Oklahoma, Rhode Island, South Dakota, Texas, Wisconsin, and West Virginia <sup>g</sup> Jurisdictions excluded from emergency department data source include California, Hawaii, Iowa, Minnesota, Oklahoma, Missouri, and Maryland <sup>h</sup> No jurisdictions were excluded from hospital admission data source

**Table 2:** Complete vaccination coverage ranges by state coverage quartile for midpointsa of Delta and Omicron predominant time periods

| Outcome | Coverage Quartile Group | Delta | Omicron |
| --- | --- | --- | --- |
| Cases<sup>b</sup> | Highest | 22-28% | 46-56% |
|  | Second Highest | 18-22% | 34-45% |
|  | Second Lowest | 14-17% | 27-34% |
|  | Lowest | 8-12% | 19-27% |
| Emergency Department Visits<sup>c</sup> | Highest | 22-29% | 45-56% |
|  | Second Highest | 17-21% | 34-43% |
|  | Second Lowest | 13-17% | 27-34% |
|  | Lowest | 8-12% | 19-27% |
| Hospital Admissions<sup>d</sup> | Highest | 22-29% | 46-56% |
|  | Second Highest | 17-22% | 35-46% |
|  | Second Lowest | 14-17% | 27-34% |
|  | Lowest | 8-13% | 19-27% |

<sup>a</sup> Midpoints of Delta and Omicron periods of predominance defined as August 22, 2021 and February 20, 2022
<sup>b</sup> Jurisdictions excluded from case data source include Alaska, Connecticut, District of Columbia, Florida, Kentucky, Hawaii, Iowa, Maryland, Missouri, Mississippi, Nebraska, New Hampshire, Oklahoma, Rhode Island, South Dakota, Texas, Wisconsin, and West Virginia
<sup>c</sup> Jurisdictions excluded from emergency department data source include California, Hawaii, Iowa, Minnesota, Oklahoma, Missouri, and Maryland
<sup>d</sup> No jurisdictions were excluded from hospital admission data source

**Figure 2:**
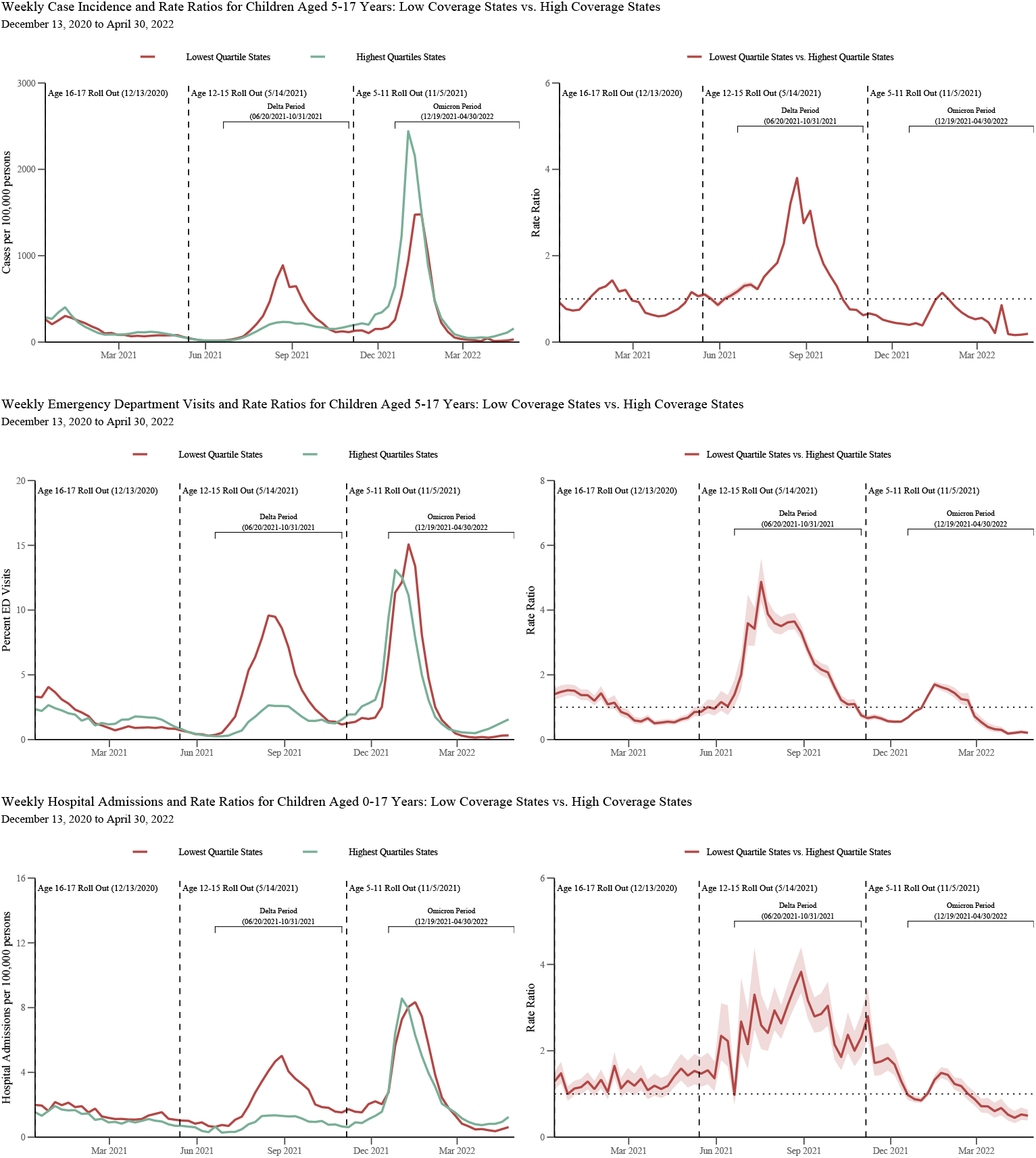
Weekly rate ratios among children aged 5–17 comparing lowest COVID-19 vaccination coverage quartile states to highest coverage quartile states by outcome: December 13, 2020–April 30, 2022 Vaccine coverage estimates for children aged 5-17 years during the first and last week of the Delta and Omicron periods of predominance in the US: (Delta: 9.21% as of the week of June 20th, 2021, 22.57% the week of October 31, 2021); (Omicron: 29.19% the week of December 19, 2021, 40.03% the week of April 24, 2022) Full vaccination coverage ranges for bottom and top quartile states as of the week of April 24, 2022: Bottom quartile: 20.14%–28.46%, Top quartile: 47.24%–59.42%

During the 3-week peak period of the Delta variant, rate ratios comparing jurisdictions in the lowest pediatric vaccination coverage quartile to the highest pediatric vaccination coverage quartile were more robust 3.19 (95% CI: 2.11–4.82) for cases, 3.41 (95% CI: 2.41–4.82) for emergency department visits, and 3.73 (95% CI: 2.70–5.14) for hospital admissions (Table 1, Figure 2). During the 3-week peak period of the Omicron variant, this association was more robust for emergency department visits: 1.28 (95% CI: 0.95–1.74).

When assessing the outcomes by age for cases and emergency department visits during the Delta period, differences were observed in the magnitude of the associations (Table 1). Children aged 12–15 and 16–17 years in the jurisdictions with the lowest vaccination coverage had a similar 2.30 (95% CI: 1.71–3.08) to 2.18 (95% CI: 1.61–2.95)-fold difference in the incidence of cases compared with jurisdictions with highest vaccination coverage, whereas this difference was 1.78-fold for children aged 5–11 years (95% CI: 1.34–2.37). Similarly, for emergency department visits, the strongest association with vaccination coverage levels by quartile were observed in children aged 12–15 (RR: 3.30, 95% CI: 2.38–4.58) and 16–17 (RR: 3.08, 95% CI: 2.28–4.17) years, versus 5–11 (RR: 2.68, 95% CI: 1.89–3.82) years. Findings were even more robust during the 3-week peak period of the Delta variant than during the full Delta period when stratifying by age group. For the Omicron period, low vaccination quartile at the state level coverage was statistically associated with lower case incidence for all age groups. Lowest vaccine coverage was significantly associated with greater emergency department visits among those aged 12–15 (RR: 1.39, 95% CI: 1.02–1.90) and 16–17 (RR: 1.34, 95% CI: 1.00–1.78) years during the 3-week peak period (Table 1).

## Discussion

In this comprehensive ecological analysis, we found that in the United States, during the period of COVID-19 Delta predominance, the rate of cases, emergency department visits, and hospital admissions were 2.00, 2.96, and 2.76 times as high, respectively, in jurisdictions with the lowest vaccination coverage compared with jurisdictions with the highest vaccination coverage. These associations were even more robust during the 3-week peak period of the Delta variant wave. This overall association was not seen during the 3-week peak period of the Omicron wave; however, children (aged 12–15 and 16–17 years) in jurisdictions with the lowest vaccination coverage had higher emergency department visits during this period.

This study mirrors the findings of recent formal vaccine effectiveness (VE) evaluations, which have found lower effectiveness during Omicron predominance compared to Delta, especially for the primary series alone^[11-13,22-23]^. During both periods, higher VE has been observed for more severe disease (e.g., hospital admissions; death), similar to our finding of a bigger impact of higher coverage on admissions and emergency department visits compared with case counts.

Consistent with our findings, the Coronavirus Disease 19 – Associated Hospitalization Surveillance Network (COVID-NET) found large increases in the rates of COVID-19-associated hospital admissions during Delta period; the monthly rate in December 2021 among unvaccinated children aged 12–17 years was six times that of vaccinated children^[20, 22-24]^.

Several possible explanations may explain the discordant findings regarding higher vaccine coverage and higher case incidence in this study and in an analysis of 68 countries and 2947 counties in the United States^[25]^. These explanations include unmeasured confounders including differences in population, vaccine products and schedules, and variant circulation during the study periods; very low vaccine coverage among children aged 5–11 years compared with adults;^[26,27]^ and waning vaccine immunity (5 months after completing a vaccine series)^[28,29]^. In addition to vaccination coverage, other factors may have affected COVID-19 incidence. Differences other than vaccine coverage (e.g., testing patterns, local and state COVID-related policies impacting school closures and mask mandates, behavior, socio-economic status, urban/rural divide) could have impacted rates of disease and were not accounted for in this analysis.

This analysis has several limitations. First, causation between pediatric vaccination coverage and outcomes cannot be inferred since this was an ecological study; pediatric vaccine effectiveness has been shown by other studies^[23]^. Second, there are limitations within data sources including lack of granular age data prior to February 2022 for hospital admissions data and data quality limitations for both case and emergency department data, leading to exclusion of some localities and jurisdictions. Nonetheless, pediatric populations in the included and excluded states were similar in regard to the distribution of their age groups, sex, and distribution by social vulnerability index^[19]^ (a measure of stressors with negative effects on communities) levels (Supplementary table 2). Third, differences in SARS-CoV-2 testing between jurisdictions with high versus low coverage could not be determined and may have biased our findings. For instance, if low coverage jurisdictions tested less, this would lower estimates of COVID-19 case incidence in these jurisdictions, and thus, potentially bias results showing less impact of the vaccine on cases. However, this would less likely affect severe outcomes (e.g., emergency department visits, and hospital admissions), where a greater impact of vaccination coverage on emergency department visits and hospital admissions was observed, when compared with cases. Finally, boosters were not approved for the youngest pediatric age group during the Delta and Omicron periods; therefore, the analysis was not able not assess the impact of additional doses after completion of the primary series.

In conclusion, during the COVID-19 Delta and Omicron waves, COVID-19 vaccination was associated with reduced COVID-19 burden on the healthcare system. Higher vaccination rates were associated with lower rates of cases, hospital admissions and emergency department visits among pediatric age groups approved to receive COVID-19 vaccine during the Delta period. A reduction in emergency department visits, but not cases or hospital admissions, was observed during the Omicron period. COVID-19 vaccines are safe and effective in preventing severe outcomes among the pediatric population^[12]^. Pediatric COVID-19 vaccinations are recommended to prevent COVID-related outcomes among children^[1-3]^.

## Data Availability

All relevant data are within the manuscript and its Supporting Information files.

